# EVALUATION OF THE PANBIO SARS-COV-2 RAPID ANTIGEN DETECTION TEST IN THE BAHAMAS

**DOI:** 10.1101/2021.07.13.21260402

**Authors:** Christoph Johnson, Kirvina Ferguson, Tavion Smith, Dashaunette Wallace, Keith McConnell, Donaldson Conserve, Indira Martin

## Abstract

Identification and isolation of persons infected with SARS-COV-2 are key mitigation strategies in the current pandemic, and rapid antigen detection tests (RADTs) offer the promise of decreased turnaround time to diagnosis when compared with gold standard RT-PCR testing. We evaluated the analytical performance of the Abbott® Panbio™ RADT on nasopharyngeal samples stored at the Ministry of Health National Reference Lab in Nassau, Bahamas. The Panbio™ demonstrated a test sensitivity of 94% and a specificity of 100% on 50 PCR negative samples and 50 samples presumed to be infectious based on having PCR cycle threshold values below 30. Additionally, in our examination of operator results there was low interpersonal variation (1%) among three blind operators and significant correlation between sample Ct value and perceived signal strength on the RADT device. However, three PCR positive samples below Ct 30 were misdiagnosed by RADT, including one sample with Ct value less than 20. These results support the use of the Panbio™ in symptomatic persons to detect active SARS-COV-2 infections, with the caveat that RADT-negative samples should be confirmed by RT-PCR.

## 1. INTRODUCTION

Rapid antigen detection tests (RADTs) represent an opportunity to expand testing for severe acute respiratory syndrome coronavirus 2 (SARS-COV-2) beyond reverse transcription polymerase chain reaction (RT-PCR), because they are easily administered in non-laboratory settings, with minimal training, and with test results available in less than an hour, compared 24-48 hours for RT-PCR testing (1). Among the many RADTs currently available on the market, the Abbott® Panbio™ nasopharyngeal swab rapid antigen test (a lateral flow immunoassay) is one of three that has been added to the World Health Organisation (WHO) Emergency Use Listing (EUL) (2) and became commercially available in the Bahamas in November 2020.

The primary goals of SARS-COV-2 diagnosis and contact tracing are to accurately identify and rapidly isolate “contagious” persons in order to limit community transmission (3). In this regard, it has emerged that samples with low viral loads/ high cycle threshold (Ct) values do not replicate in viral culture, which has been used as a surrogate for contagiousness or infectivity. Significantly, several studies have demonstrated negligible cultivability of samples with N or E gene Ct values above 27-35 (with the wide range probably due to variability in the RT-PCR and viral culture assays used) (4-9). However, while it has been established that RADT test sensitivity decreases at around Ct 25-30 (10-18), recent reports find that RADT results correlate more to viral cultivability than do RT-PCR results, supporting the postulate that the RADT detects active SARS-COV-2 infections, and that there is minimal clinical significance to their reduced sensitivity at higher Ct values compared to gold standard RT-PCR testing (19-21).

We sought in this study to assess the analytical performance of the Panbio™ in a selection of 100 frozen stored samples at the Ministry of Health National Reference Lab (NRL) in the Bahamas, including 50 RT-PCR negative samples and 50 RT-PCR positive samples with presumed infectious quantities of virus as garnered by their Ct value (<30). The objective of this analysis was to specifically assess the ability of trained operators using the Panbio™ RADT to detect SARS-COV-2 active infections in symptomatic, presumed contagious individuals with high viral loads.

## 2. METHODS

Samples obtained from symptomatic persons meeting the clinical case definition for SARS-COV-2 infection (3) from community primary public health clinics on islands throughout the Bahamas archipelago are routinely sent to NRL. 100 de-identified retrospective samples stored frozen in viral transport medium (VTM) at the NRL were selected using the following criteria: a) 50 PCR positive samples from symptomatic persons (E gene Ct value <30) and b) 50 PCR negative samples from persons with ILI symptomatology. These samples were selected from the month of November 2020, during the ‘second wave’ of the covid-19 epidemic in the Bahamas.

150ul of each thawed VTM sample was combined with 150ul of the Abbott® extraction buffer (dilution ratio 1:1) and 130ul of the mixed solution was dispensed onto the Abbott® Panbio™ rapid antigen test device as per manufacturer’s product insert (22). Results were recorded in 15-20 minutes by three independent testers. Samples were previously identified as SARS-COV-2 positive by detection of the E gene using the Charite (Berlin) RT-PCR protocol (23) on the ABI QuantStudio 6 Flex at the NRL.

RADT performance and results interpretation was performed by three independent operators concurrently performing blind testing; operators were asked to score their results as negative (score of 0), weakly detected-1, or strongly detected-2. In the case of discordance among the three operators, the consensus result was taken. Sensitivity was measured by the following formula: number of true positives/ number of true positives + number of false negatives. Specificity was measured by the formula: number of true negatives/ number of true negatives + number of false positives. Limit of detection (LOD) relative to E gene Ct value was estimated by comparison of Ct values of serial dilutions of a known PCR positive sample with the RADT result. The Spearman’s rank coefficient was calculated for operator-ranked results and graphs constructed in Graphpad Prism Version 9.0.2.

## 3. RESULTS

### 3.1. Study population

50 SARS-COV-2 PCR-negative samples and 50 PCR positive frozen stored VTM samples with Ct values <30-derived from 8 islands within the Bahamas archipelago: Grand Bahama, Eleuthera, Exuma, Bimini, Andros, New Providence, Abaco, and Inagua-were selected for the evaluation. Median age was 38.5 and 39 years old in positive and negative sample groups, respectively. 58% of positive samples and 36% of negative samples were female. Median Ct value in the PCR positive sample group was 22.91 (Table 1). Samples were collected during the month of November 2020, during the peak of the ‘second wave’ of the SARS-COV-2 epidemic in the Bahamas.

**Table 1:** Study population demographics

|  | RT-PCR<br>Positive<br>samples, n | (% of<br>total) | RT-PCR<br>Negative<br>samples, n | (% of<br>total) |
| --- | --- | --- | --- | --- |
| **Total no. of samples** | 50 | (100) | 50 | (100) |
| **Gender** |  |  |  |  |
| Female | 29 | (58) | 18 | (36) |
| Male | 21 | (42) | 32 | (64) |
| **Island of Origin** |  |  |  |  |
| Grand Bahama | 10 | (20) | 9 | (18) |
| Eleuthera | 4 | (8) | 5 | (10) |
| Exuma | 24 | (48) | 1 | (2) |
| Bimini | 7 | (14) | 2 | (4) |
| Andros | 1 | (2) | 1 | (2) |
| New Providence | 3 | (6) | 31 | (62) |
| Abaco | 1 | (2) | 0 | (0) |
| Inagua | 0 | (0) | 1 | (2) |
| **Median Age (Years)** | 38.5 |  | 39 |  |
| **CT value (median)** | 22.91 |  | N/A |  |

### 3.2. Abbott® Panbio™ RADT performance

RADT performance characteristics are summarised in Table 2. 50 of 50 (100%) PCR negative samples were correctly identified by the device. However, 3 of 50 PCR positive samples received false negative results. Distribution of Ct values among false negative samples is shown in Figure 1; two were relatively high Ct/ low viral load samples (Ct value 28.44 and 29.49, respectively) and one was a low Ct/high viral load sample (Ct value 19.37). Overall, this resulted in a product sensitivity of 94% and specificity of 100% in presumed infectious samples.

**Table 2:**
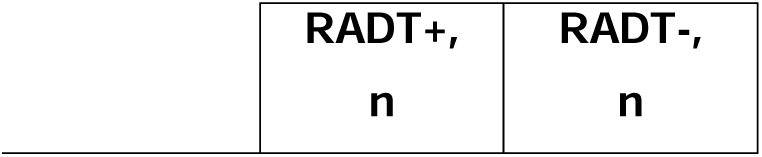
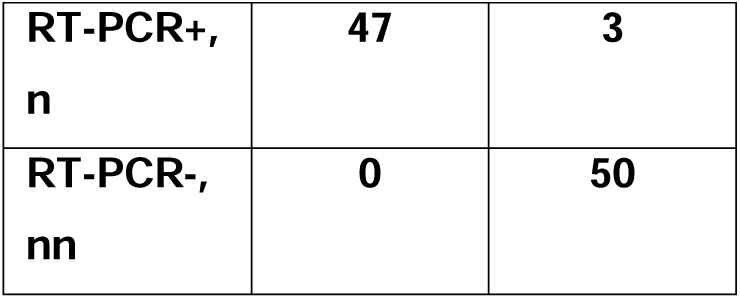
Summary of test performance characteristics

| RADT+,<br>n | RADT-,<br>n |
| --- | --- |

|  |  |  |
| --- | --- | --- |
| RT-PCR+,<br>n | 47 | 3 |
| RT-PCR-,<br>nn | 0 | 50 |

**Figure 1A:**
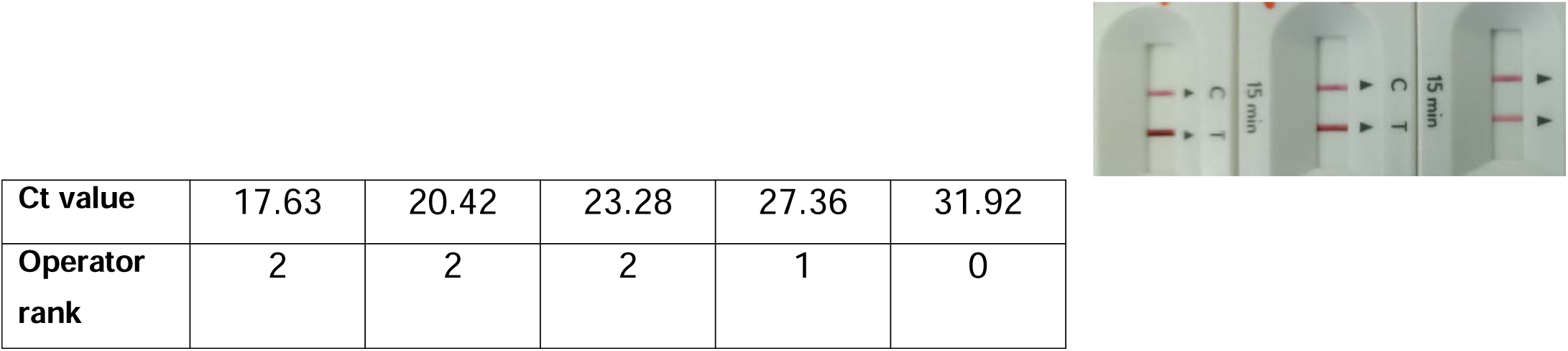
Image of Abbott® Panbio™ detection of range of sample serial dilutions and Ct values with corresponding RADT test result ranked by operators as strong (2), weak (1) and negative (0)

**Figure 1B:**
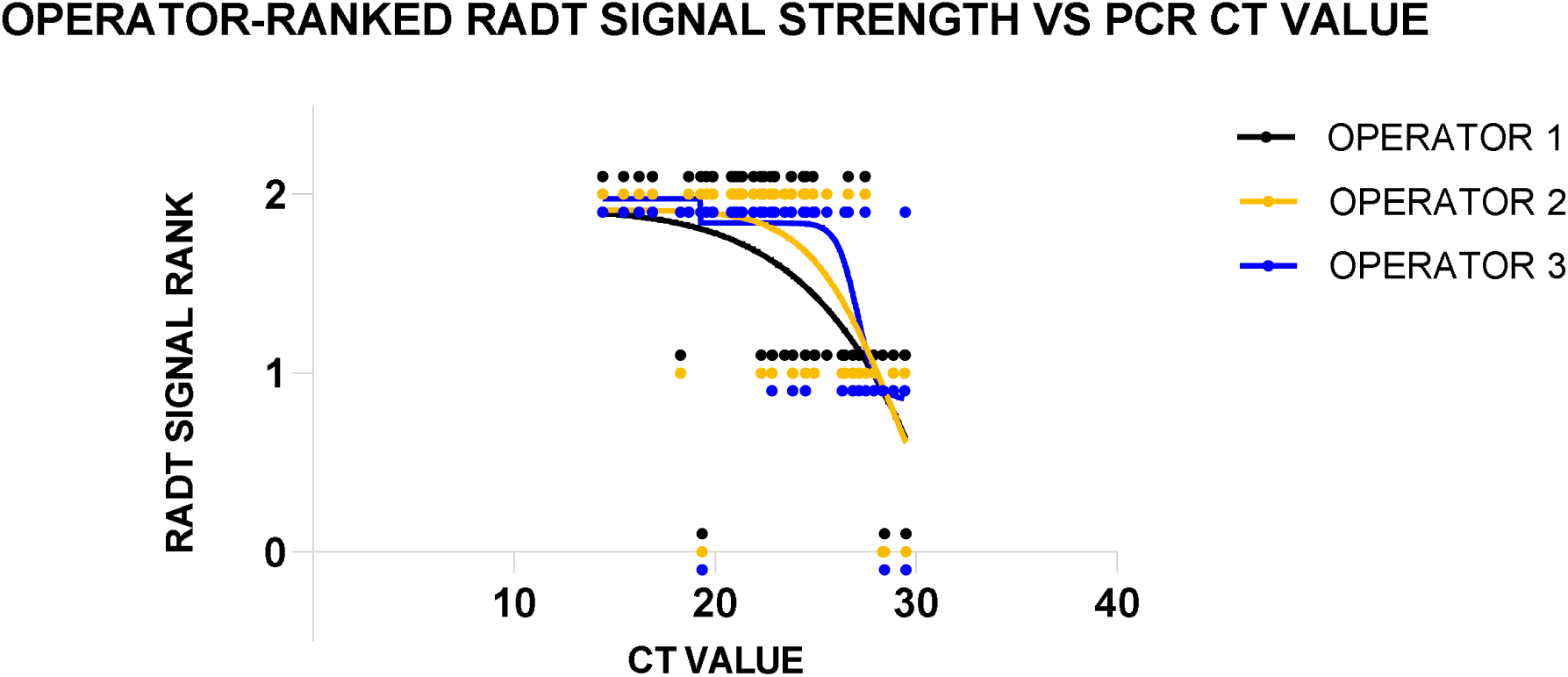
Relationship between operator-ranked RADT signal strength and Ct value of 50 PCR positive samples, with curve of best fit for each of 3 operators (r_s_ = -0.59, - 0.57 and -0.53, respectively (p=0.001, 95% CI= -0.29 to -0.75)).

### 3.3. Limits of detection (LODs)

LOD for signal strength=1 (weak) was between E gene approximate Ct values 27 and 32, and for signal strength=2 (strong) was between approximate Ct 23 and 27 (Figure 2A). Operator-ranked signal strength (i.e. 0, 1, 2) of the 50 PCR positive samples was moderately inversely correlated with sample Ct value (r_s_ = -0.59, -0.57 and -0.59, p=0.001 for each of the three operators)-see Figure 2B.

### 3.4. Inter-personal variation

Of 3 independent operators reading qualitative results (i.e., RADT positive or negative) on blinded samples, overall there was 1% discordance (described as a dissenting result among the three submitted), due to one of the three operators misdiagnosing a PCR positive sample (Ct 28.35) as negative on the Abbott® RADT.

## 4. DISCUSSION

Overall, the Panbio™ RADT performed well in our study samples derived from symptomatic individuals throughout the Bahamas archipelago, with a sensitivity and specificity of 94% and 100% respectively in samples with Ct value <30. Notwithstanding this, and in common with previous studies, we found that high Ct value is not the sole determinant of false negative test results on the Panbio™ RADT, since one sample with Ct value <20 was given a false negative result by all three test operators. This phenomenon has been seen in other settings where the Panbio™ RADT produced false negatives in a few of these high viral load samples (10,13). Interestingly, this issue of missing some low Ct samples seems to be a general feature of RADTs, as revealed in a recent head-to-head comparison between 4 RADTs and an automated antigen test, in which the latter was the only among those evaluated that did not exhibit this phenomenon (13). Though an apparently relatively rare and a minor contributor to overall test performance in this and previous studies, false negative results in low Ct/high viral load samples may be significant in real-world contexts in which missing active infections can lead to preventable transmission events (although the clinical relevance of this remains to be seen, since RADT results correlate better with viral cultivability than PCR results (19-21))

In our opinion, this observation reveals an important limitation to use of RADTs for mass screening activities and also lends strong support to following an algorithm in which symptomatic RADT negative persons are re-tested (by RADT or preferably RT-PCR), as is recommended by the WHO (1). Indeed, previous evaluations of RADT accuracy in asymptomatic groups, such as children (24) and household contacts of known cases (25) have reported low sensitivity of the Panbio™ in these populations, probably due to low viral load (high Ct value).

In previous analyses the Panbio™ test LOD has been described as correspondent to a Ct value of between 24 and 29 (10-18). Our LOD results using sample serial dilution were consistent with these published LODs, since we estimate the LOD as between approximate Ct 28 and 32, as there was a weak line detected in a sample with Ct 28 but no line at Ct 32 (Figure 2A). In addition, blinded operator-ranked results of RADT signal intensity were significantly inversely correlated in the nonparametric analysis to sample Ct value (Figure 2B). Notably, however, samples as low as Ct 23 were concordantly ranked as weak (score=1) by all operators (Figure 2B). Therefore, despite a concordance of 99% among 3 independent operators, and the fact that the single discordant result recorded by one of the operators was reported on a sample with a relatively high Ct value (28.35) and faint RADT band, there remains the possibility that samples with Ct values <25 could produce “weak” signal strength on the RADT and be subject to interpersonal variation leading to misdiagnosis of potentially infectious samples.

The impact of interpersonal variation in Panbio™ test result interpretation can be minimised by measures such as implementation of two-reader policy, or device automation. In addition, expansion of quality assurance services for SARS-COV-2 antigen testing, such as proficiency testing, can help to identify sources of error in test administration and results interpretation particularly in borderline/faint samples (26).

In totality, the present analysis supports the utility of the Panbio™ RADT within the prescribed parameters of early symptomatic infection, demonstrating a high sensitivity in presumed infectious samples. However, use of this test must be tempered with the understanding that it may (on relatively rare occasions) misdiagnose low viral load/high Ct value samples and that these samples may produce faint test lines on the rapid test device leading to false negative results due to variability in results interpretation. In addition to this, there are instances where low Ct value/high viral load samples produce weak RADT signal strength and/or false negative results for yet unknown reasons. Thus, while RADT is a useful tool to enhance access to SARS-COV-2 testing, this data also supports cautious rollout, limited to detection of cases in the early symptomatic phase, and supported by quality oversight and RT-PCR confirmation of RADT-negative symptomatic cases.

## Data Availability

All data not contained in the manuscript are available upon request

## ETHICAL APPROVAL

Ethics approval for this study was granted by the Bahamas National Covid-19 Ethics Committee, April 2021.

## ACKNOWLEDGEMENTS

We gratefully acknowledge Dr Tyneil Cargill (Dynamic Health Services) and Dr Esther de Gourville (immediate past Pan American Health Organisation representative for the Bahamas).

## REFERENCES

1. Antigen-detection in the diagnosis of SARS-CoV-2 infection using rapid immunoassays: interim guidance, 11 September 2020. World Health Organization. https://apps.who.int/iris/handle/10665/334253. License: CC BY-NC-SA 3.0 IGO

2. WHO Emergency Use Listing for In vitro diagnostics (IVDs) Detecting SARS-CoV-2 https://extranet.who.int/pqweb/sites/default/files/documents/210126_eul_sars_cov2_product_list.pdf

3. Public health surveillance for COVID-19: interim guidance: Interim guidance, 16 December 2020. World Health Organization. https://www.who.int/publications/i/item/who-2019-nCoV-surveillanceguidance-2020.8 License: CC BY-NC-SA 3.0 IGO

4. Viral RNA load as determined by cell culture as a management tool for discharge of SARS-CoV-2 patients from infectious disease wards. La Scola B, Le Bideau M, Andreani J, Hoang VT, Grimaldier C, Colson P, Gautret P, Raoult D. (2020) Eur J Clin Microbiol Infect Dis. 39(6):1059–1061.

5. An approach to lifting self-isolation for health care workers with prolonged shedding of SARS-CoV-2 RNA. Laferl H, Kelani H, Seitz T, Holzer B, Zimpernik I, Steinrigl A, Schmoll F, Wenisch C, Allerberger F. (2021) Infection 49(1):95–101.

6. Duration of infectiousness and correlation with RT-PCR cycle threshold values in cases of COVID-19, England, January to May 2020. Singanayagam A, Patel M, Charlett A, Lopez Bernal J, Saliba V, Ellis J, Ladhani S, Zambon M, Gopal R. (2020) Euro Surveill. 25(32):2001483

7. Viral cultures for COVID-19 infectious potential assessment - a systematic review. Jefferson T, Spencer EA, Brassey J, Heneghan C. (2020) Clin Infect Dis. 3:ciaa1764.

8. Predicting infectious SARS-CoV-2 from diagnostic samples. Bullard J, Dust K, Funk D, Strong JE, Alexander D, Garnett L, Boodman C, Bello A, Hedley A, Schiffman Z, Doan K, Bastien N, Li Y, Van Caeseele PG, Poliquin G. (2020) Clin Infect Dis. 22:ciaa638

9. Repeat COVID-19 Molecular Testing: Correlation of SARS-CoV-2 Culture with Molecular Assays and Cycle Thresholds. Gniazdowski V, Morris CP, Wohl S, Mehoke T, Ramakrishnan S, Thielen P, Powell H, Smith B, Armstrong DT, Herrera M, Reifsnyder C, Sevdali M, Carroll KC, Pekosz A, Mostafa HH. (2020) Clin Infect Dis. Oct 27:ciaa1616.

10. Evaluation of the Panbio COVID-19 Rapid Antigen Detection Test Device for the Screening of Patients with COVID-19. Fenollar F, Bouam A, Ballouche M, Fuster L, Prudent E, Colson P, Tissot-Dupont H, Million M, Drancourt M, Raoult D, Fournier PE. (2021) J Clin Microbiol. 59(2):e02589–20.

11. Field evaluation of a rapid antigen test (Panbio™ COVID-19 Ag Rapid Test Device) for COVID-19 diagnosis in primary healthcare centres. Albert E, Torres I, Bueno F, Huntley D, Molla E, Fernández-Fuentes MÁ, Martínez M, Poujois S, Forqué L, Valdivia A, Solano de la Asunción C, Ferrer J, Colomina J, Navarro D. (2020) Clin Microbiol Infect. Nov 13:S1198-743X(20)30697-2.

12. Evaluating the clinical utility and sensitivity of SARS-CoV-2 antigen testing in relation to RT-PCR Ct values. Lanser L, Bellmann-Weiler R, Öttl KW, Huber L, Griesmacher A, Theurl I, Weiss G. (2020) Infection. Nov 13:1–3.

13. Head-to-Head Comparison of Rapid and Automated Antigen Detection Tests for the Diagnosis of SARS-CoV-2 Infection. Favresse J, Gillot C, Oliveira M, Cadrobbi J, Elsen M, Eucher C, Laffineur K, Rosseels C, Van Eeckhoudt S, Nicolas JB, Morimont L, Dogné JM, Douxfils J. (2021) J Clin Med. 10(2):265.

14. Panbio antigen rapid test is reliable to diagnose SARS-CoV-2 infection in the first 7 days after the onset of symptoms. Linares M, Pérez-Tanoira R, Carrero A, Romanyk J, Pérez-García F, Gómez-Herruz P, Arroyo T, Cuadros J. (2020) J Clin Virol. Dec;133:104659.

15. Evaluation of rapid antigen test for detection of SARS-CoV-2 virus. Mak GC, Cheng PK, Lau SS, Wong KK, Lau CS, Lam ET, Chan RC, Tsang DN. (2020) J Clin Virol. Aug;129:104500.

16. Multicenter evaluation of the Panbio™ COVID-19 rapid antigen-detection test for the diagnosis of SARS-CoV-2 infection. Merino P, Guinea J, Muñoz-Gallego I, González-Donapetry P, Galán JC, Antona N, Cilla G, Hernáez-Crespo S, Díaz-de Tuesta JL, Gual-de Torrella A, González-Romo F, Escribano P, Sánchez-Castellano MÁ, Sota-Busselo M, Delgado-Iribarren A, García J, Cantón R, Muñoz P, Folgueira MD, Cuenca-Estrella M, Oteo-Iglesias J; Spanish Panbio™ COVID-19 validation group. (2021) Clin Microbiol Infect. Feb 16:S1198-743X(21)00076-8.

17. Real-life validation of the Panbio COVID-19 antigen rapid test (Abbott) in community-dwelling subjects with symptoms of potential SARS-CoV-2 infection. Gremmels H, Winkel BMF, Schuurman R, Rosingh A, Rigter NAM, Rodriguez O, Ubijaan J, Wensing AMJ, Bonten MJM, Hofstra LM. (2021) EClinicalMedicine. Jan;31:100677.

18. Field evaluation of a rapid antigen test (Panbio COVID-19 Ag Rapid Test Device) for COVID-19 diagnosis in primary healthcare centres. Albert E, Torres I, Bueno F, Huntley D, Molla E, Fernández-Fuentes MÁ, Martínez M, Poujois S, Forqué L, Valdivia A, Solano de la Asunción C, Ferrer J, Colomina J, Navarro D. (2020) Clin Microbiol Infect. Nov 13:S1198-743X(20)30697-2.

19. Evaluation of a SARS-CoV-2 rapid antigen test: Potential to help reduce community spread? Toptan T, Eckermann L, Pfeiffer AE, Hoehl S, Ciesek S, Drosten C, Corman VM. (2021) J Clin Virol. 135:104713.

20. Antigen-Based Testing but Not Real-Time Polymerase Chain Reaction Correlates With Severe Acute Respiratory Syndrome Coronavirus 2 Viral Culture. Pekosz A, Parvu V, Li M, Andrews JC, Manabe YC, Kodsi S, Gary DS, Roger-Dalbert C, Leitch J, Cooper CK. (2021) Clin Infect Dis. Jan 20:ciaa1706.

21. The Comparative Clinical Performance of Four SARS-CoV-2 Rapid Antigen Tests and Their Correlation to Infectivity In Vitro. Kohmer N, Toptan T, Pallas C, Karaca O, Pfeiffer A, Westhaus S, Widera M, Berger A, Hoehl S, Kammel M, Ciesek S, Rabenau HF. (2021) J Clin Med. 10(2):328

22. Abbott Panbio COVID-19 Ag Rapid Test Device Nasal Procedure Instructions For Use (Multilingual) https://www.globalpointofcare.abbott/en/product-details/panbio-covid-19-ag-antigen-test.html

23. Detection of 2019 novel coronavirus (2019-nCoV) by real-time RT-PCR. Corman VM, Landt O, Kaiser M, Molenkamp R, Meijer A, Chu DK, Bleicker T, Brünink S, Schneider J, Schmidt ML, Mulders DG, Haagmans BL, van der Veer B, van den Brink S, Wijsman L, Goderski G, Romette JL, Ellis J, Zambon M, Peiris M, Goossens H, Reusken C, Koopmans MP, Drosten C. (2020) Euro Surveill. 25(3):2000045.

24. Think of the Children: Evaluation of SARS-CoV-2 Rapid Antigen Test in Pediatric Population. González-Donapetry P, García-Clemente P, Bloise I, García-Sánchez C, Sánchez Castellano MÁ, Romero MP, Gutiérrez Arroyo A, Mingorance J, de Ceano-Vivas La Calle M, García-Rodriguez J; SARS-CoV-2 Working Group (2021). Pediatr Infect Dis J. Feb 17.

25. Evaluation of a rapid antigen test (Panbio COVID-19 Ag rapid test device) for SARS-CoV-2 detection in asymptomatic close contacts of COVID-19 patients. Torres I, Poujois S, Albert E, Colomina J, Navarro D (2021). Clin Microbiol Infect. Jan 6:S1198-743X(20)30782-5.

26. SARS-CoV-2 antigen-detecting rapid diagnostic tests: an implementation guide. Geneva: World Health Organization; 2020. Licence: CC BY-NC-SA 3.0 IGO.

